# Prophylactic Para Aortic Irradiation vs Pelvic Radiotherapy in Pelvic node-positive Carcinoma Cervix in the setting of concurrent chemoradiation: A phase II Open-label Multi centric Randomized Controlled Trial (PRO-PARA)

**DOI:** 10.1101/2024.04.16.24305717

**Authors:** Tapesh Bhattacharyya, Santam Chakraborty, Bhavana Rai, Shirley Lewis, Srinivas Gowda, Anurupa Mahata, Samar Mandal, Gaurav Trivedi, Sreekripa Rao, Sarath Shyan

**Author notes:** Funding: Extramural funding from Indian Council of Medical Research. **Role of sponsor**:Extramural funding has been obtained from the Indian Council for Medical Research. The ICMR has no role in the study design as the collection, management, analysis, and interpretation of data; writing of the report; and the decision to submit the report for publication.

## Abstract

**Background:** EMBRACE and Retro EMBRACE studies have shown that excellent local control and pelvic control could be achieved with concurrent chemoradiation and MRI-based brachytherapy in carcinoma cervix. Now para aortic nodal failure and distant metastasis are the predominant modes of failure. Paraaortic nodal failure rates are higher in pelvic node-positive cases as compared to pelvic node-negative cases as demonstrated in EMBRACE studies. The current study aims to find out the benefit of adding prophylactic para-aortic node irradiation in patients of carcinoma cervix who have involved pelvic nodes on volumetric imaging.

**Method:** This will be a two-arm, parallel group, phase II open-label multicenter randomized controlled trial. Patients will be enrolled in a phase II trial where the primary endpoint will be demonstration of reduction in the risk of para-aortic recurrence.If the primary endpoint is met, a phase III trial will be initiated using the same trial design and intervention. Patients in arm A(control arm) will receive pelvic radiotherapy covering the common iliac nodes with Intensity Modulated Radiotherapy (IMRT) to a dose of 45 Gy/25 fractions over five weeks. Radiologically involved lymph nodes will be boosted to a dose of 55 Gy/25 fractions with simultaneous integrated boost(SIB).Patients in arm B (Experimental arm)will receive pelvic and elective para-aortic radiotherapy up to the lower border of the renal vein (IMRT) to dose of 45 Gy/25 fractions over five weeks.

Radiologically involved lymph nodes will be boosted to a dose of 55 Gy/25 fractions with simultaneous integrated boost(SIB). Concurrent chemotherapy with cisplatin 40mg/m2 weekly will be given during external beam radiotherapy in both the arms. After completion of concurrent chemoradiation, high dose rate (HDR) intracavitary or intracavitary +interstitial brachytherapy will be performed in both the arms.

With a one sided type I error of 5% and a power of 80%, a total of 9 para-aortic recurrences is required to demonstrated that addition of prophylactic para-aortic nodal recurrence reduces the the risk of a para-aortic nodal failure from 10% in the control arm to 2% or less in the test arm. Without a substantial reduction in the risk of para-aortic nodal failure, an improvement in overall survival cannot be expected. 224 patients will need to be accrued over a period of 2 years with a minimum follow up of 12 months to demonstrate this number of para-aortic nodal recurrences for the phase II trial.

For the current study we will assume that the 5 year overall survival is 70% in the control arm and that use of prophylactic EFRT will translate into an absolute improvement of 9% in the overall survival. This implies that the test arm will have a 5 year overall survival of about 79%. This corresponds to a hazard ratio of 0.75 which is a conservative estimate of the possible relative benefit of extended field radiotherapy. With a two sided type I error of 5% and a power of 80%, a total of 143 events is required to demonstrate an improvement in the overall survival corresponding to the hazard ratio of 0.75. This would need a total accrual duration of 5 years, and a minimum follow up duration of 4 years (such that the total trial duration of 9 years), a total of 462 patients (equal allocation) need to be accrued. Assuming a 15% loss to follow up, a total sample size of 530 patients is needed corresponding to an annual accrual of 106 patients..

**Discussion:** This trial will demonstrate the efficacy of prophylactic para aortic radiation in pelvic node positive carcinoma cervix. It also gives an opportunity to standardize and assess the quality-assurance radiotherapy practices in carcinoma cervix across multiple premier institutes of the nation at the same time.The safety of this intervention in advanced pelvic node-positive disease requiring prophylactic para aortic radiation will be established.

**Trial Registration:** The trial has been registered at the Clinical Trial Registry of India (CTRI) vide registration number: CTRI/2023/08/057075(30th August 2023)

## Introduction

### Background and Rationale

Definitive concurrent chemoradiation is the standard of care for locally advanced carcinoma cervix [1–6].EMBRACE and Retro EMBRACE studies have shown excellent treatment outcomes for patients with locally advanced cervical cancer (LACC) treated with concomitant chemoradiation (CTRT) and MRI (Magnetic Resonance Imaging)-based image-guided brachytherapy (IGBT) [7,8]. Advances in IGBT resulted in 3 years of local/pelvic control and overall survival of 86-97% and 65-87% respectively [7,8].In the setting of excellent local control now achievable with conformal external beam radiotherapy and MR-IGBT, para-aortic nodal failure and distant failures are the dominant causes of treatment failure.

In EMBRACE study, patients without pathological nodes at diagnosis have significantly fewer paraaortic failures compared to those with node-positive disease (4% versus 11%). Pathologic nodes at diagnosis are mainly located in the lower pelvis. Nodal failures however are more often seen in the paraaortic region. About 40% of all failures were reported outside the treatment targets[7].

The initial results of the prophylactic extended field radiotherapy encompassing the paraaortic region in carcinoma cervix have shown conflicting results.Radiation Therapy Oncology Group (RTOG) 79-20 study showed overall survival (OS) advantage and benefit in reducing distant metastasis with elective para aortic nodal coverage compared to traditional whole pelvic radiotherapy in the absence of chemotherapy [9,10]. The study enrolled 367 patients and demonstrated that 10 year overall survival improved from 44% to 54% with the use of extended field radiotherapy [10]. On the other hand, results from the European Organization for Research and Treatment of Cancer (EORTC) showed only a potential decrease in isolated PA nodal failures with an extended field technique, but no differences in local control, distant metastases, or overall survival [11]

Peters et al have presented an analysis of the factors predicting the occurrence of nodal failure in EMBRACE I observational study.In this study of the 1338 patients, 152 patients had nodal failure and 104 patients had para-aortic nodal failure [12]. Elective para-aortic nodal radiation was associated with a reduction in the risk of para-aortic nodal failure with hazard ratio of 0.53 (95% CI 0.28 - 1.00) on multivariate analysis.

Modern radiotherapy techniques utilizing intensity-modulated radiation therapy (IMRT), and image-guided radiation therapy (IGRT) have shown reduced acute and late toxicities [13,14]. While 2D techniques of extended field radiotherapy have been historically associated with significant bowel toxicity, modern intensity modulated radiotherapy combined with concurrent chemotherapy is relatively better tolerated [15]. In a recent propensity score matched comparison of extended field radiotherapy versus pelvic radiotherapy alone, Wang et al demonstrated a significant benefit from EFRT in nodal relapse, disease free survival and overall survival.This benefit was demonstrated irrespective of the number of nodes involved in the pelvis[16]. However, no randomized studies have assessed the outcomes between whole pelvic versus extended field techniques utilizing these techniques with concurrent chemotherapy. This background has prompted us to plan a prospective phase II randomized controlled trial comparing prophylactic para-aortic radiation to standard pelvic radiotherapy in patients with a high risk of para-aortic nodal recurrence.

#### Background and Rationale: Choice of comparators

EMBRACE [7] and Retro EMBRACE [8] studies have shown that excellent local control and pelvic control could be achieved with concurrent chemoradiation and MRI-based brachytherapy in carcinoma cervix. Now paraaortic nodal failure and distant metastasis are the predominant modes of failure. Paraaortic nodal failure rates are higher in pelvic node-positive cases as compared to pelvic node-negative cases as demonstrated in EMBRACE studies. The current study aims to find out the benefit of adding prophylactic para-aortic node irradiation in patients of carcinoma cervix who have involved pelvic nodes on volumetric imaging.

#### Objectives

We hypothesize that prophylactic para aortic radiation along with concurrent chemotherapy along with modern day image based brachytherapy will reduce the chances of para aortic failure rate as compared to concurrent chemoradiation with pelvic radiotherapy alone.

The primary endpoint is to compare the para-aortic nodal recurrence rate between the two arms (time point : 3 years).

The secondary endpoints are:

1. To compare the progression-free survival (PFS) between the two arms (Time point: 3 years)
2. To compare the distant recurrence rate between the two arms. (Time point: 3 years)
3. To compare the pelvic nodal recurrence rate between the two arms (Time point :3 years)
4. To compare the local failure rate between the two arms (Time point : 3 years)
5. To compare the cumulative incidence of acute and late toxicities between the two arms. Toxicity will be reported using CTCAE(Common terminology criteria for adverse events) version.5.0.
6. To compare the global quality of life, symptom experience, body image scale scores between the two arms at the end of external beam radiotherapy, and at 6 months, 1 year, 2 year and 3 years. EORTC(European Organisation for Research and Treatment) QLQC30 and CX24 modules will be used for evaluating the quality of life of patients.

## Methods and Analysis

### Trial design

This is a multicentric, open-label, parallel group, phase II randomized controlled trial designed to demonstrate that use of prophylactic extended field radiotherapy can improve the para-aortic nodal control as compared to standard pelvic radiotherapy. Stratified randomization will be performed with a 1:1 allocation. Patients will be randomized through a centralized randomization unit. Treatment allocation will be done using stratified randomization using the following stratification factors

1. Center of the study
2. Neoadjuvant chemotherapy planned or not
3. Involvement of common iliac nodes: Present or Absent
4. Histology: Squamous or non-squamous

If the phase II trial demonstrates absence of futility, a phase III trial will be started with overall survival as the end-point. Given the direct effect of prophylactic para-aortic nodal radiation is to reduce the risk in para-aortic nodal failure, a demonstrable improvement in the overall survival can be expected only if this is reduced substantially. Therefore the phase II component will be designed to determine if the prophylactic para-aortic nodal radiation is futile with respect to para-aortic nodal recurrence rate. If the futility is disproven then the trial will continue to accrue till the recruitment target of the phase III trial is met to test for an improvement in overall survival.Trial schema is displayed in **Figure 1**.

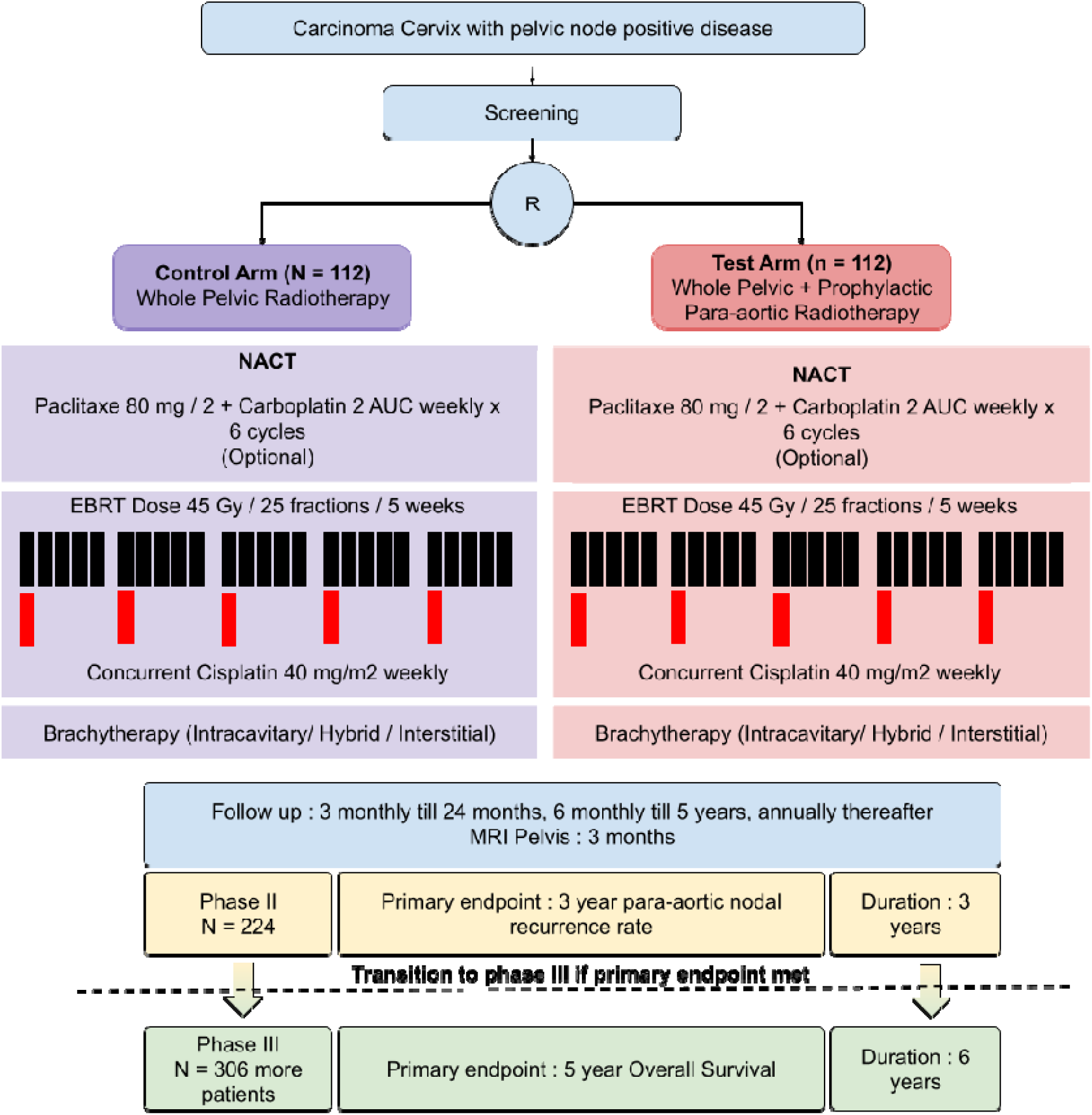

### Study setting

The trial is currently supported by extramural funding from the ICMR(Indian Council of Medical Research).The study has been initiated at the Department of Radiation Oncology **at Tata Medical Center, Kolkata.Site initiation has been done at PGIMER Chandigarh and**

**Kasturba Medical College, Manipal.** Additional centers may be enrolled after adequate quality assurance. All centers selected will be high volume centers with a known track record of delivering high quality intensity modulated radiotherapy for patients.

### Eligibility criteria

The selection criteria has been designed in such a way that the cohort is enriched with patients who have a higher risk of para-aortic nodal recurrence in absence of para-aortic nodal radiation and closely aligns with the selection criteria for prophylactic extended field radiation in the EMBRACE II cohort study.

The following are the **inclusion criteria** for this trial:

1. Age : 18 years or more.
2. Epithelial cervical cancer: Squamous cell carcinoma, adenocarcinoma, and adenosquamous cell carcinoma confirmed by pathology at Tata Medical Center or PGI Chandigarh, Kasturba Medical College, Manipal or any laboratory accredited by EQAS(External Quality Assurance Scheme).
3. Patients with pelvic node positive disease. Pelvic node positivity can be determined based on pathological evaluation or in the presence of radiological features as:

a. Short axis dimension of 1 cm or higher.
b. Presence of 0.5 - 1 cm node with features suspicious of metastatic involvement (any of the below):

i. Abnormal FDG PET uptake as determined by a qualified nuclear medicine physician
ii. Diffusion restriction in MRI
iii. T2 signal hyperintensity in MRI
iv. Rounded shape
v. Irregular margins suggestive of extracapsular extension.
vi. Presence of central necrosis
4. Highest level of the radiologically involved pelvic node is below 1 cm from the aortic bifurcation
5. Cisplatin based chemotherapy can be given:

a. Adequate bone marrow function (white blood cells ≥3·0×10⁹/L, platelets ≥100×10⁹/L)
b. Liver function (bilirubin ≤1·5×upper normal limit [UNL], aspartate aminotransferase and alanine aminotransferase ≤2·5×UNL)
c. Kidney function (creatinine clearance >60 mL per min calculated according to Cockroft and Gault or > 50 mL per min DTPA)
6. Eastern Cooperative Oncology Group (ECOG) score 0-2.
7. Women of childbearing age must have a pregnancy test (serum or urine) within 7 days before enrollment, and the result is negative, and they are willing to use appropriate contraceptive methods during the test.

The primary **exclusion criteria** are as follows:

1. Patients with paraaortic node involvement on CT scan (documented by imaging or pathology)
2. Stage IVA or Stage IVB disease
3. Presence of pathologically confirmed inguinal nodes.
4. Patients requiring elective coverage of the inguinal region.
5. Uncontrolled severe infections
6. Epithelial cancers of other types, neuro-endocrine tumors, melanomas, sarcomas, germ cell tumors.
7. Patients with synchronous malignancies who need treatment for another tumor.
8. Patients with prior history of malignancy excluding cervical intraepithelial neoplasia or basal cell carcinoma of the skin.
9. Liver cirrhosis, decompensated liver disease, chronic renal insufficiency, and renal failure
10. Presence of uncontrolled or severe comorbidities which preclude safe concurrent chemoradiation e.g Myocardial infarction, severe arrhythmia, and grade 2 or more congestive heart failure (NYHA classification).
11. Prior history of renal transplantation
12. Horseshoe kidney or single kidney
13. History of severe allergic reaction to platinum-containing chemotherapy drugs
14. Patients with a history of autoimmune diseases are known to increase the risk of late radiation induced adverse effects especially Systemic Lupus Erythematosus.

### Pretreatment Evaluation

All patients should have the following pretreatment evaluation for the study:

1. Clinical assessment including a medical history and examination of the patient. The primary tumor size, vaginal involvement and parametrial extension should be noted in the records.
2. Biopsy should be obtained from the cervix for documentation of invasive cancer. Biopsy report with carcinoma in situ is not sufficient for the study. A repeat biopsy should be obtained or reviewed at the trial center.
3. Complete hemogram: Hemoglobin, total leucocyte count, platelet count and differential leucocyte count.
4. Renal function test : Serum Urea and Creatinine
5. Electrolytes : Serum Sodium, Potassium, Magnesium and Calcium
6. Liver function test
7. HIV(Human Immunodeficiency Virus), HBV(Hepatitis B Virus) and HCV(Hepatitis C Virus) status
8. CEMRI(Contrast enhanced Magnetic Resonance Imaging) of the Pelvis with standard MRI protocol
9. Contrast enhanced CT (Computed Tomography)scan of Thorax and Abdomen or FDG (Fluorodeoxyglucose) /PET (Positron emission tomography)CT

These investigations are performed as a standard for all new patients presenting with carcinoma cervix at the participating centers.

### Study Interventions

Eligible patients would be randomized in equal proportions between PRT (Pelvic Radiotherapy) and EFRT(Extended Field Radiotherapy). Radiation therapy will be delivered to the following volumes:

### Control arm:(Arm A)

Patients in arm A will receive pelvic radiotherapy only to a dose of 45 Gy/25 fractions over five weeks. Pelvic radiotherapy will include the primary clinical target volume (CTV) along with the common iliac, external iliac, internal iliac, presacral and obturator nodal CTV. Radiologically involved lymph nodes will be boosted to a dose of 55Gy /25 fractions with simultaneous integrated boost (SIB). Concurrent chemotherapy with cisplatin 40mg/m2 weekly will be given during external beam radiotherapy. After completion of concurrent chemoradiation, a high dose rate(HDR) brachytherapy with 7Gyx4 sessions of intracavitary or intracavitary with interstitial brachytherapy will be performed. The targeted cumulative dose to 90% of the volume of the high-risk clinical target volume (D90 HR CTV) should be between 80-85Gy.

### Test arm:(Arm B)

Patients in arm B will receive pelvic and elective para-aortic radiotherapy to a dose of 45Gy /25 fractions over five weeks. Para-aortic radiotherapy will be delivered to a clinical target volume which extends to the lower border of the left renal vein. Inferiorly it extends into the pelvic nodal CTV. Radiologically involved lymph nodes will be boosted to a dose of 55Gy /25 fractions with simultaneous integrated boost(SIB). Concurrent chemotherapy with cisplatin 40mg/m2 weekly will be given during external beam radiotherapy. After completion of concurrent chemoradiation, a high dose rate (HDR) brachytherapy of 7Gy X 4 sessions of intracavitary or intracavitary +interstitial brachytherapy will be performed. The linear quadratic equivalent dose at 2 Gy to the high-risk clinical target volume(HR CTV) D90 should be more than ≥80 Gy depending on HRCTV volume.

### Radiotherapy

#### Timing

Radiotherapy should be started within 3 weeks of randomization if neoadjuvant chemotherapy is not used. In patients receiving neoadjuvant chemotherapy, radiotherapy shoould be started within 7 - 10 weeks post randomization. Concurrent chemotherapy should be initiated within 3 days of starting radiotherapy.

## Equipment

Megavoltage equipment capable of delivering static intensity modulation with a multileaf collimator or dynamic intensity modulation (using a multileaf collimator or tomotherapy) is required. For all patients the allowed energies are 6 - 15 MV photons. For isocentric linear accelerators mounted on a C-arm gantry the Source to Source Distance (SSD) should be 100 cm.

## Localization, Simulation and positioning

The patient preparation procedures will remain uniform and will not be dependent on the arm allocated. All patients will undergo CT simulation. Use of an immobilizing device is allowed if pre-specified in the institutional protocol. Patients can be positioned supine or prone depending on the institutional protocol, but the same positioning should be used for all patients. Patients will be scanned in the helical mode with 2.5 mm slices. CT scans will be obtained from diaphragm to the mid thigh. A uniform bladder filling protocol is required to be pre-specified. Additionally centers may pre-specify any protocol used for ensuring reproducible rectal filling. Centers may choose either of the methods to account for the uterine motion:

1. A margin based approach with a uniform or differential margin is given around the uterus to account for the motion and positioning changes with bladder or rectal filling. The margin may be anisotropic but should be pre-specified in the institutional protocol.
2. An approach based on generating an ITV based on the summation of the extent of uterine motion on the full and empty bladder scans. In such a case, the empty bladder scan should be obtained after the patient has passed urine. The full bladder scan will be obtained with contrast after the patient has been instructed to drink 500 ml of water and wait for 45 minutes.

## Target volume delineation

If the institute opts for a ITV based approach, then the full bladder and empty bladder scans will be rigidly registered with the volume of interest encompassing the pelvis. The result of the rigid registration would be visually verified to ensure that bones in the pelvis are appropriately matched. If the institute opts for a margin based approach then this step is not needed.

### Primary CTV

Primary CTV will encompass the gross disease and the entire cervix, uterus, and adnexa (including the ovaries). Vaginal extent of the disease will determine the length of vagina to be included in the primary CTV. Typically at least 2 cm of the normal vagina beyond the clinically or radiologically demonstrated disease should be included in the CTV. The only exception will be when oophoropexy will be done for ovarian preservation. In that case, the adnexa will not be encompassed in the CTV. CTV primary should include parametrium which will extend superiorly up to the peritoneal reflection (guided by the presence of the round ligament or the sigmoid colon). Anteriorly the boundary will be the posterior wall of the bladder or the posterior border of the external iliac vessel. Laterally it will extend to the pelvic vessels. Inferiorly the volume will be bounded by the urogenital diaphragm. Mesorectum will be included if clinically there is heavy involvement of mesorectal space or there are perirectal nodes. Additionally, for patients with mesorectal nodes or mesorectal involvement by the disease, the entire mesorectum will be included in the CTV. For institutes opting for a ITV based approach, the primary CTV will be delineated in full and empty bladder CT scans and added together to generate a composite ITV.

### Nodal CTV

Pelvic vessels will be delineated (both artery and vein) including the common iliac, external iliac and the visceral branches of the internal iliac vessels. The following expansions will be given around the vessels to generate the nodal CTV using the following steps:

1. 7 mm isotropic margin would be given first around the vessels.
2. The generated volume will be edited such that it is edited away from the muscles, bone, bowel and bladder.

The **Table 1** shows the CTV volume boundaries

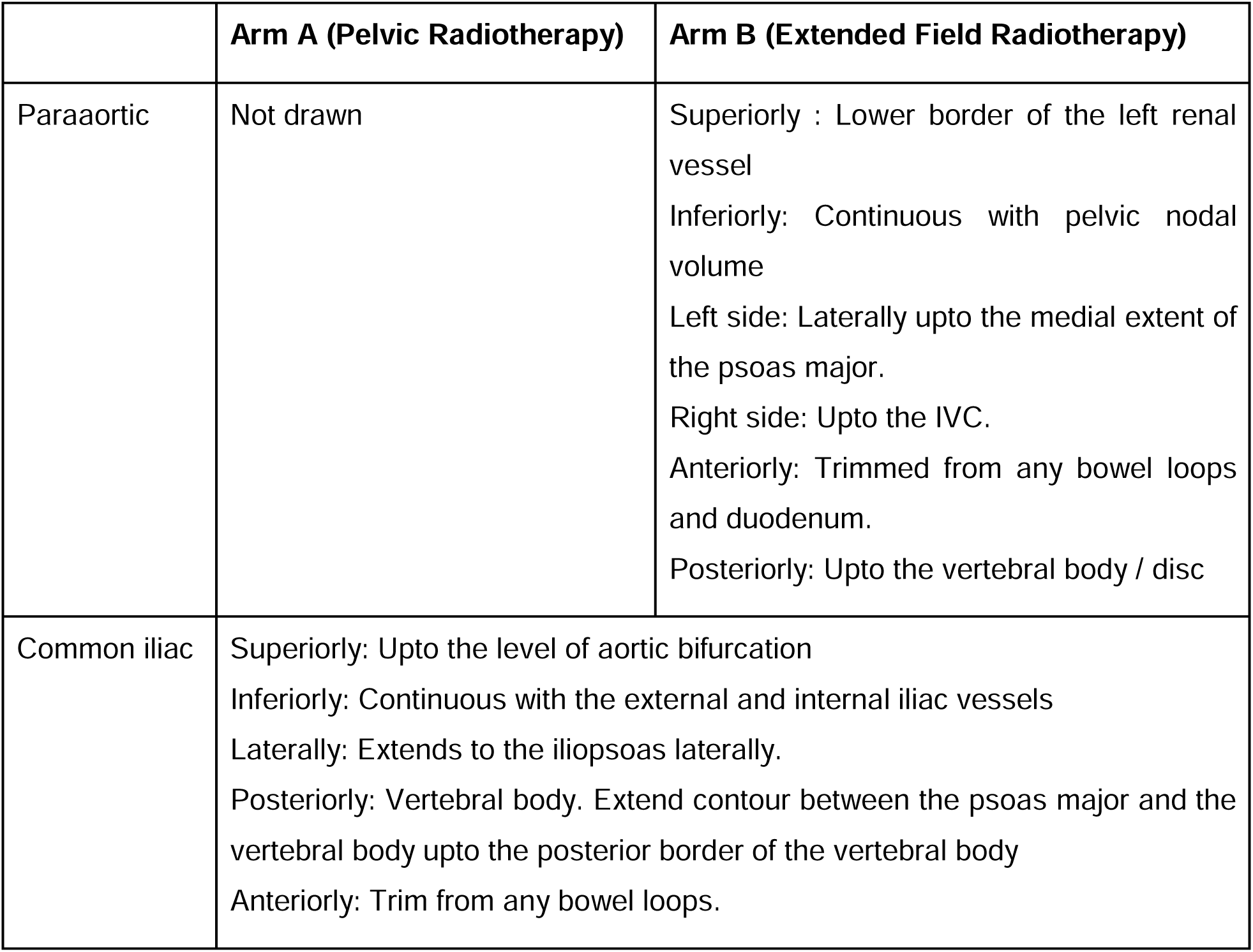
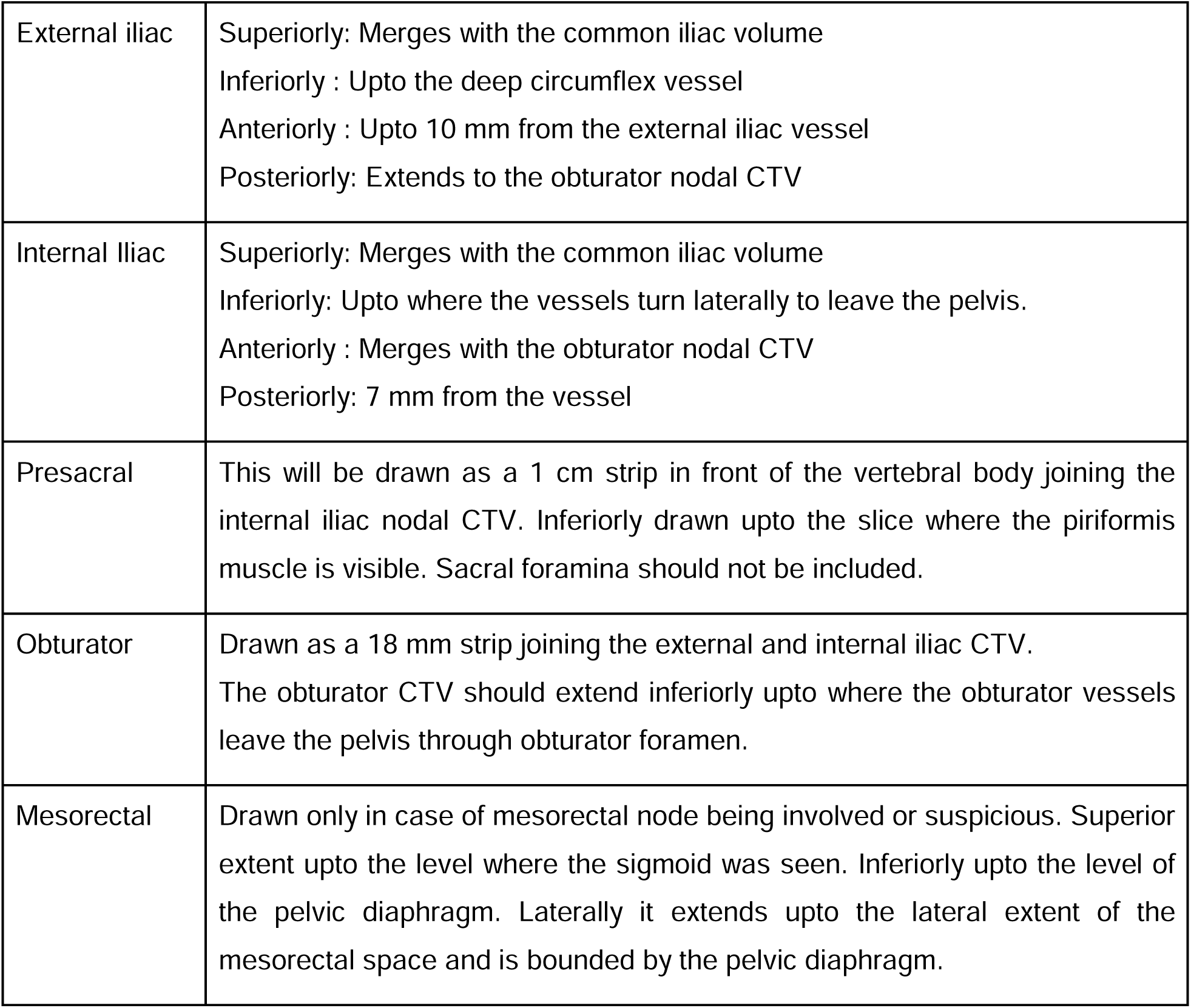

### PTV(Planning Target Volume)

Will be generated based on the institutional protocol. Usually the primary CTV and nodal CTV are joined together and a geometric expansion will be applied to the total CTV. The PTV will not be trimmed from any structure unless it extends beyond the body contour in which case it will be trimmed back by 5 mm from the body surface to allow correct dose calculation. In order to specify this the institute should have data of 50 consecutive patients treated with IMRT in the machine(s) in which the treatment is to be delivered. The planning target volume will be calculated using the Van Herk formula.

A high dose boost PTV will be generated, in addition, by giving the same margin to the gross nodal volume as being used for the primary PTV.

### Critical Structures

The following will be delineated for all patients:

1. Femoral heads: Will be delineated separately for the left and right side. Will include the femoral head, neck, greater trochanter and the portion of femur up until the lower border of the lesser trochanter.
2. Bowel Bag: Delineated as a single structure that includes the entire external contour of the bowel. Superiorly it should extend to at least 2 cm above the planning target volume
3. Bone marrow : Pelvic, sacrum and lumbar vertebra will be delineated as a single structure. Femoral heads will not be included in the bone marrow contour.
4. Rectum: The rectum will be delineated as a single structure from the anal verge to the sigmoid colon (where the rectum turns anteriorly)
5. Bladder: Will be drawn as a single structure to include the entire bladder.

In patients who receive EFRT the following structures will be delineated in addition:

1. Kidney: Will be delineated separately for the left and right side. The entire kidney will be included in the contour along with the renal pelvis.
2. Duodenum: Will be drawn as a single structure to include the 1st, 2nd, 3rd and 4th parts of the duodenum.
3. Pancreas: Will be drawn as a single structure to include the head, body and tail of pancreas along with the uncinate process.
4. Spinal canal: Will be drawn as a single structure extending upto the level of the lower border of the body of the L3 vertebra.

Additional organs at risk and supporting structures may be delineated as per institutional practice but the doses to the above structures are to be mandatorily reported.

### Dose fractionation

For both arms, the external beam radiotherapy dose is 45 Gy in 25 fractions delivered over 5 weeks treating 5 fractions in a day and a single fraction each day. Twice daily treatments are not allowed unless mandated to compensate for unplanned gaps in treatment. Breaks or planned gaps in treatment are not allowed except to allow for interdigitating brachytherapy applications.

### Dose Specification

No point based dose prescription is needed though a reference point may be generated for the planning process. No plan normalization is allowed and dose should be prescribed to the 100% isodose. The following will be the dose specification for the volumes:

1. Whole PTV : 45 Gy in 25 fractions over 5 weeks.
2. Boost PTV: 10 Gy in 25 fractions over 5 weeks delivered as a simultaneous integrated boost (SIB) to a total dose of 55 Gy in 25 fractions over 5 weeks.

## Treatment Planning

As inverse planned intensity modulated radiotherapy is mandated in this protocol, the field arrangements, collimator alignment and other beam parameters will be determined by the planning approach used in each institute. Use of non-coplanar fields is allowed. However if field junctions are needed for adequate coverage the junctional dose should be measured in at least 5 patients as a part of the quality assurance process. Both volumetric arc and fixed field approaches for intensity modulation are allowed. However, each institute should specify the planning technique to be adopted in the process document for the study. Irrespective of the planning technique used, the following dose constraints should be met as planning objectives. Physicists are free to create dummy structures to support plan optimization. Additionally, MU objectives and Normal Tissue Avoidance objectives available in the treatment planning systems can be used as long as the planning strategy is pre-specified. Institutes will be encouraged to designate machines for which treatment planning can be done for this trial. As this will be a volumetric prescription, the ICRU 84 recommendations for dose reporting should be followed.

**Table 2** is showing the dose constraints to be followed for the trial. Mandatory constraints should be met in all cases. In case these are not met, it would be considered as a protocol violation. These doses are with respect to a prescribed dose of 45 Gy for the whole PTV and 55 Gy to the boosted node.

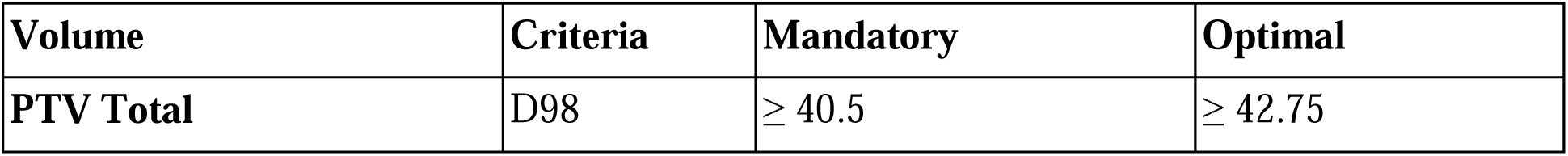
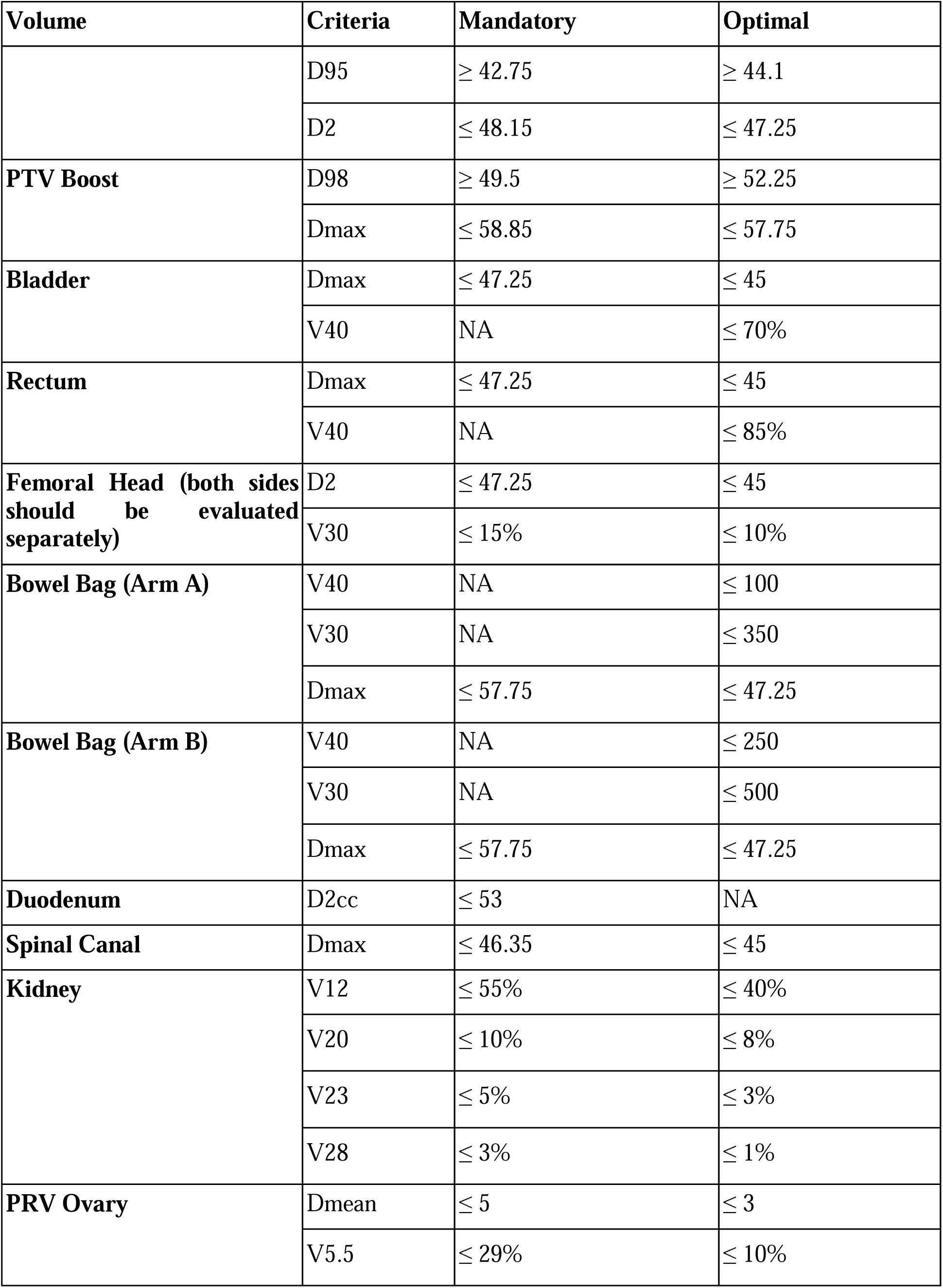
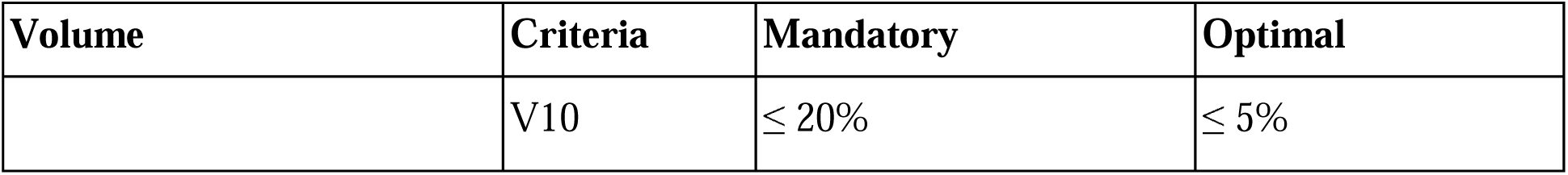

| Volume | Criteria | Mandatory | Optimal |
| --- | --- | --- | --- |
| PTV Total | D98 | $\geq 40.5$ | $\geq 42.75$ |

| **Volume** | **Criteria** | **Mandatory** | **Optimal** |
| --- | --- | --- | --- |
| | D95 | $\geq 42.75$ | $\geq 44.1$ |
| | D2 | $\leq 48.15$ | $\leq 47.25$ |
| **PTV Boost** | D98 | $\geq 49.5$ | $\geq 52.25$ |
| | Dmax | $\leq 58.85$ | $\leq 57.75$ |
| **Bladder** | Dmax | $\leq 47.25$ | $\leq 45$ |
| | V40 | NA | $\leq 70\%$ |
| **Rectum** | Dmax | $\leq 47.25$ | $\leq 45$ |
| | V40 | NA | $\leq 85\%$ |
| **Femoral Head (both sides should be evaluated separately)** | D2 | $\leq 47.25$ | $\leq 45$ |
| | V30 | $\leq 15\%$ | $\leq 10\%$ |
| **Bowel Bag (Arm A)** | V40 | NA | $\leq 100$ |
| | V30 | NA | $\leq 350$ |
| | Dmax | $\leq 57.75$ | $\leq 47.25$ |
| **Bowel Bag (Arm B)** | V40 | NA | $\leq 250$ |
| | V30 | NA | $\leq 500$ |
| | Dmax | $\leq 57.75$ | $\leq 47.25$ |
| **Duodenum** | D2cc | $\leq 53$ | NA |
| **Spinal Canal** | Dmax | $\leq 46.35$ | $\leq 45$ |
| **Kidney** | V12 | $\leq 55\%$ | $\leq 40\%$ |
| | V20 | $\leq 10\%$ | $\leq 8\%$ |
| | V23 | $\leq 5\%$ | $\leq 3\%$ |
| | V28 | $\leq 3\%$ | $\leq 1\%$ |
| **PRV Ovary** | Dmean | $\leq 5$ | $\leq 3$ |
| | V5.5 | $\leq 29\%$ | $\leq 10\%$ |
| | V10 | $\leq 20\%$ | $\leq 5\%$ |

Bone marrow dose constraints are likely to be different between the two techniques due to the larger volume of marrow treated with extended field radiotherapy. While dose recording of the bone marrow doses will be done, an explicit constraint on the bone marrow dose is not being specified as evidence based dose volume constraints for bone marrow irradiation in extended field radiotherapy are not well established. The data from this trial may help in establishing an appropriate dose volume constraint in the future.

### Beam Energy

Most patients will be planned with 6 MV photons. Use of flattening filter free 6 MV and 10 MV photons is allowed if adequately commissioned for use with IMRT.

### Plan Quality Assurance

As a trial which mandates Intensity Modulated Radiotherapy, per patient plan quality assurance is necessary. For all patients, a verification plan will be created on a phantom. Point dose verification will be done at isocenter with the help of an ion chamber. Variation within 3% of the prescribed dose will be considered acceptable. Additional fluence verification with the help of additional ion chambers at other points, ion chamber / diode arrays and films are allowed.

### Intervention Modification

#### Administering Neoadjuvant Chemotherapy

Use of neoadjuvant chemotherapy as used in the INTERLACE trial protocol [17] is allowed for eligible patients in both arms. In case this is used the same should be specified in the process document for the institute. The regimen allowed for this will be weekly Paclitaxel (80 mg / m2) with weekly Carboplatin (2 AUC) for a maximum of six cycles. If neoadjuvant chemotherapy is used, patients should be started on radiotherapy 1 week after the last cycle of chemotherapy. Hence treatment planning should be done in the 2nd to 3rd week of chemotherapy. In all such cases, the pre-chemotherapy target volume should be used for treatment planning (specially in case of good response to vaginal or uterine disease). Response imaging is not needed after neoadjuvant chemotherapy. Concurrent chemoradiation protocol will remain consistent. It is important that randomization is done before neoadjuvant chemotherapy.

Treatment should not be interrupted unless the patient has particularly severe Grade IV toxicity. In particular external beam radiotherapy treatment should not be interrupted for thrombocytopenia unless counts go below 20,000. In such cases a daily manual platelet count may be mandated before treatment. Use of platelet transfusions is allowed to maintain the radiation dose intensity. Similarly radiation should not be held for neutropenia and use of growth factor support is allowed. However in case of severe Grade IV hematological toxicity the investigator may decide to omit extended field radiotherapy. All such cases should be clearly recorded as a protocol violation, and the dose of radiation delivered as EFRT should be recorded in the case record form.

#### Interventions: adherence

During treatment every patient will be reviewed in review clinic where their physical and blood parameters will be checked and physical signs and symptoms will be assessed. It will enable us to intervention adherence and improve the compliance to treatment.

#### Interventions: concomitant care

The subjects of this trial will not be able to participate in another parallel trial during the trial time period. No specific contraindications to any medication.

### Outcomes

#### Primary Outcome

The primary outcome measure is the proportion of patients who have para aortic recurrences at 3 years.

1. Para-aortic recurrence rate (PARR): Defined as the cumulative incidence of para-aortic nodal recurrence. Para-aortic nodal recurrence is any nodal recurrence noted in the lateral aortic, retroaortic, inter-aortocaval, retrocaval, paracaval, pre-aortic, and pre-caval region between the level of the left renal vein and the aortic bifurcation. The recurrence rate will be calculated by the Fine Gray method with death being considered as a competing event.3 year cumulative incidence of para-aortic recurrence between the two arms will be reported with corresponding 95% confidence intervals.

#### Secondary Outcomes

1. Progression Free survival (PFS): Defined as the time interval between the date of randomization and the date of disease progression (any disease progression) or death due to any cause. Second malignant neoplasms will not be considered as events. In patients who have residual disease, and undergo salvage surgery, the date of documentation of residual disease will be considered as the date of event. In patients with suspected residual disease, but who do not undergo surgery, the date when the disease progressed locally or elsewhere will be considered as the date of progression. The 3 year PFS will be reported for both arms along with the corresponding 95% confidence intervals.
2. Distant metastasis free survival (DMFS): Defined as the time between the date of randomization and the date of distant metastases (sites include any part of the body except locoregional recurrence, para-aortic and pelvic nodes). Thus mediastinal nodes, retrocrural nodes and inguinal nodal metastases would be considered as distant metastases. As far as possible an attempt should be made to obtain a biopsy confirmation from the metastatic site unless clinically not indicated. 3 year DMFS will be reported for both arms along with the 95% confidence intervals.
3. Pelvic nodal recurrence rate (PNRR): Defined as the cumulative incidence of pelvic nodal recurrence. Pelvic nodal recurrence is any nodal recurrence noted in the pelvis below the aortic bifurcation and above the level of the inguinal nodes. Recurrence in presacral, mesorectal, parametrial and obturator nodes will be considered as a recurrence. The recurrence rate will be calculated by the Fine Gray method with death being considered as a competing event. 3 year cumulative incidence of pelvic nodal recurrence between the two arms will be reported with corresponding 95% confidence intervals.
4. Local recurrence rate (LRR): Defined as the cumulative incidence of local failure at the cervix, uterus, upper half of the vagina or parametrium. In order to qualify as a local recurrence, the disease should have a biopsy or a strong clinical suspicion. The recurrence rate will be calculated by the Fine Gray method with death being considered as a competing event. 3 year cumulative incidence of local recurrence between the two arms will be reported with corresponding 95% confidence intervals.
5. Acute toxicity: Acute toxicities will be defined as per the CTCAE v 5.0 criteria. Acute toxicities will be considered as any toxicity related to chemotherapy and radiation which are observed during treatment and upto 90 days post completion of treatment. For each toxicity, the proportion of patients with Grade II, III and IV CTCAE events will be reported for the two arms.
6. Late Toxicity: Late toxicities will be also defined as per the CTCAE v 5.0 criteria. Late toxicities are any radiation or chemotherapy related toxicities which are observed after 90 days of completion of treatment. For each toxicity we will be reporting the cumulative proportion of patients experiencing a grade II, III and IV event. Additionally the time to first occurrence of Grade III or more toxicity for rectal proctitis, bladder cystitis, bowel obstruction, bowel perforation, bowel fistula, ureteric or vesical fistula, vesicovaginal or rectovaginal fistula, pelvic insufficiency fracture, avascular necrosis of the femur, duodenal bleeding and second malignancies will be estimated
7. Quality of life: Quality of life will be recorded using the EORTC QLQ C30 and CX24 questionnaires in the quality of life substudy. The mean scores of the global quality of life score (from the EORTC QLQ C30 questionnaire), symptom experience (CX 24 questionnaire), lymphedema (CX 24) and body image scale (CX 24 questionnaire) at end of treatment, 6 month, 12 month, 24 month and 36 month post treatment will be reported. Additional analysis of PROMs will be conducted as per the substudy protocol.

#### Participant timeline

##### On treatment assessment

Patients will be followed weekly during chemoradiation with routine clinical review and toxicity grading (CTCAE 5.0).

### Quality of Life Assessment

Quality of life assessments will be performed for patients enrolled in the quality of life substudy using the European Organization for Research and Treatment of Cancer (EORTC) quality of life questionnaire 30 (QLQ C30) and the Cervix 24 (EORTC CX 24) questionnaire. Bengali, Punjabi and Hindi translations will be made available so that patients can self-report quality of life assessments. Assessments will be performed at baseline, end of radiotherapy and then at 6, 12, 24 and 36 months after radiotherapy. Additional assessments at interim follow up visits may also be conducted. Paper and electronic administration is allowed. Additionally, quality of life data can be collected through telephonic interviews if the patient follow-up cannot be maintained physically Participant timeline diagram is displayed in **Table 3**.

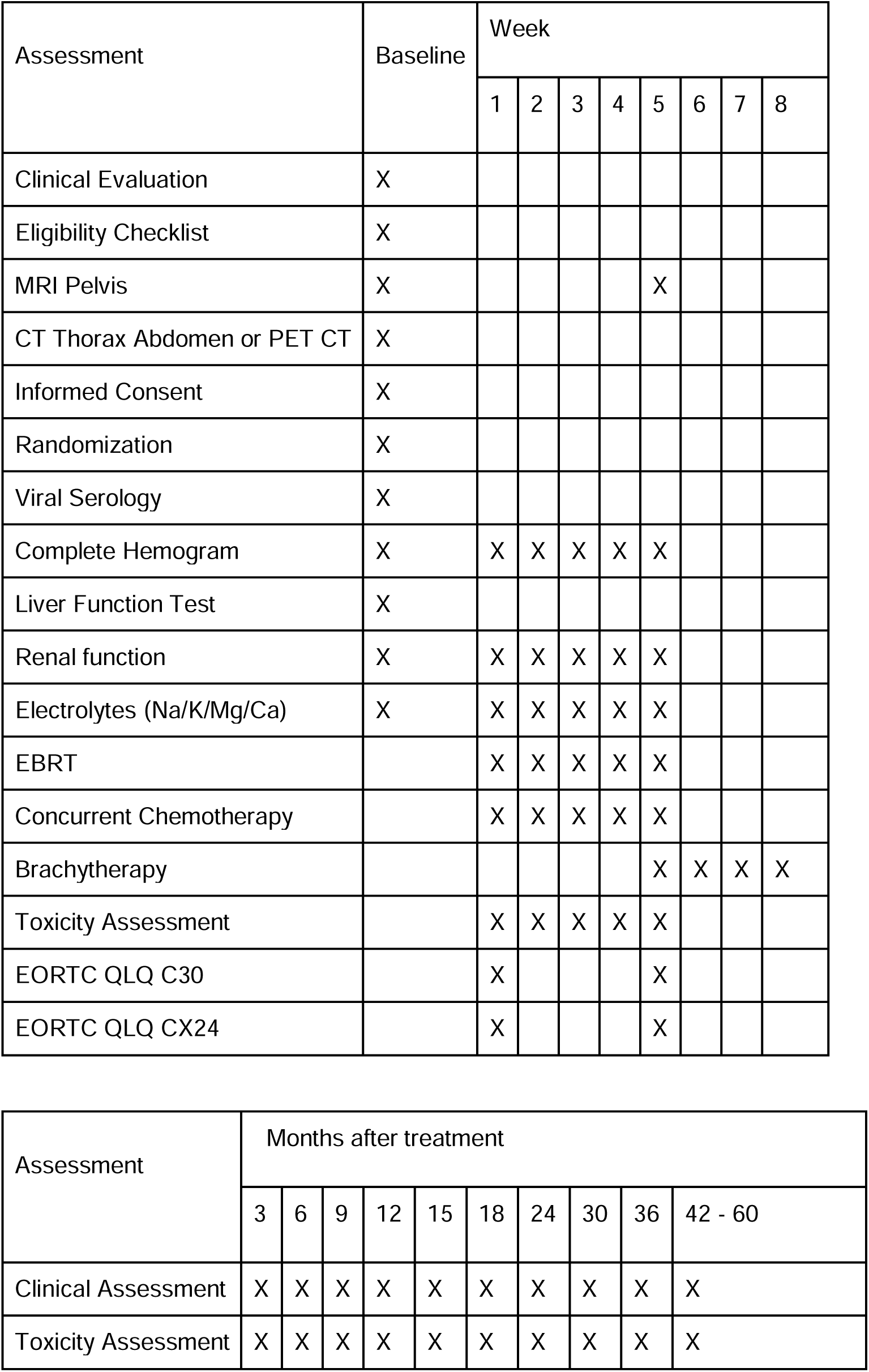
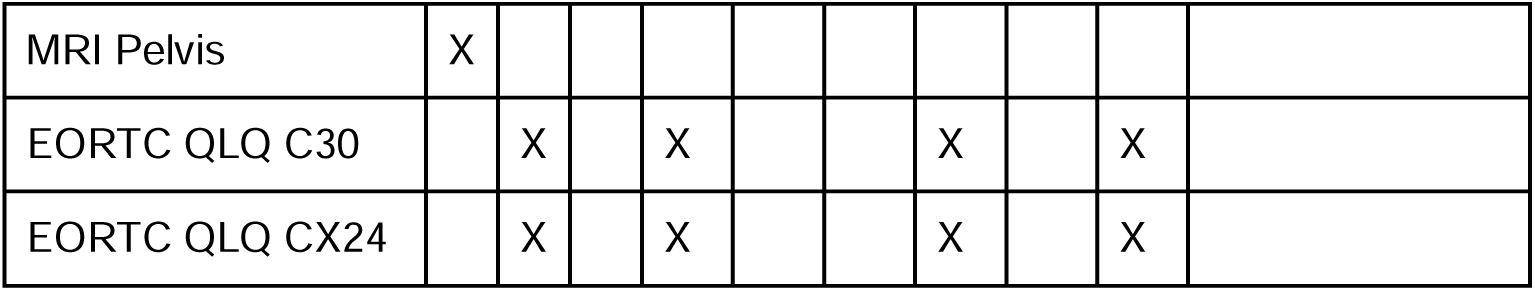

Participant timeline: Table 3
| Assessment | Baseline | Week |  |  |  |  |  |  |  |
| --- | --- | --- | --- | --- | --- | --- | --- | --- | --- |
|  |  | 1 | 2 | 3 | 4 | 5 | 6 | 7 | 8 |
| Clinical Evaluation | X |  |  |  |  |  |  |  |  |
| Eligibility Checklist | X |  |  |  |  |  |  |  |  |
| MRI Pelvis | X |  |  |  |  | X |  |  |  |
| CT Thorax Abdomen or PET CT | X |  |  |  |  |  |  |  |  |
| Informed Consent | X |  |  |  |  |  |  |  |  |
| Randomization | X |  |  |  |  |  |  |  |  |
| Viral Serology | X |  |  |  |  |  |  |  |  |
| Complete Hemogram | X | X | X | X | X | X |  |  |  |
| Liver Function Test | X |  |  |  |  |  |  |  |  |
| Renal function | X | X | X | X | X | X |  |  |  |
| Electrolytes (Na/K/Mg/Ca) | X | X | X | X | X | X |  |  |  |
| EBRT |  | X | X | X | X | X |  |  |  |
| Concurrent Chemotherapy |  | X | X | X | X | X |  |  |  |
| Brachytherapy |  |  |  |  |  | X | X | X | X |
| Toxicity Assessment |  | X | X | X | X | X |  |  |  |
| EORTC QLQ C30 |  | X |  |  |  | X |  |  |  |
| EORTC QLQ CX24 |  | X |  |  |  | X |  |  |  |
| MRI Pelvis | X |  |  |  |  |  |  |  |  |
| EORTC QLQ C30 |  | X |  | X |  |  | X |  | X |
| EORTC QLQ CX24 |  | X |  | X |  |  | X |  | X |

**Participant timeline: Table 3**

#### Follow up

After completion of treatment patients will be reviewed 3 monthly for 2 years, and then 6 monthly till 5 years. After 5 years an annual follow up will be recommended. Each patient is planned to complete the study treatment unless disease progression occurs or toxicity (CTCAE 5.0) prohibits further therapy. At each follow up patient will undergo a clinical evaluation to document the disease status and document late toxicities. Imaging is not required in these patients but symptom directed imaging can be ordered as required. The patient follow up schedule will be maintained in the RED Cap database and patients will be contacted at regular intervals to ensure adherence to follow up visits .In case the patient is unable to come for a physical follow up telemedicine follow-ups are allowed. The end of study visit will be completed in case of patient death, withdrawal of consent or at 3 years after the last patient is enrolled in the trial.

#### Statistical Analysis and Sample size

For the phase II component of the trial, we chose the primary endpoint of para-aortic nodal recurrence rate expressed as the cumulative incidence of para-aortic nodal recurrence at 3 years. From previous studies, we know that the para-aortic nodal recurrence occurs in 10 - 15% patients. Given that the population of patients planned for accrual in this trial are expected to have a higher risk of para-aortic nodal recurrence it is reasonable to assume that about 10% of the patients in the control arm can be expected to have a para-aortic nodal recurrence at 3 years. The immediate benefit expected from para-aortic nodal radiation is a reduction in para-aortic nodal recurrence. Hence unless a robust reduction in para-aortic nodal recurrence is demonstrated, the trial will not accrue further as a difference in overall survival is unlikely to be demonstrated. The trial will not continue further unless the para-aortic nodal recurrence rate is 2% or lesser in the arm receiving prophylactic para-aortic nodal radiotherapy. As shown in our literature review it is not unreasonable to expect this number of para-aortic recurrences in our trial population at 3 years.

A total of 9 para-aortic nodal recurrences will provide a power of 80% to detect this difference with a one-sided Type I error rate of 5%. Note that in this case we are looking at a futility end point (i.e. if the reduction in the incidence of para-aortic nodal recurrence is lower the trial will not proceed to phase III), one sided testing is justified. The required number of events is expected to occur after 224 patients have been accrued and observed over a minimum period of 12 months. Going by the projected accrual rate in this trial we expect that it will take us 2 years to complete the accrual for the phase II trial. Also as we will not analyze the overall survival as a part of this phase II analysis adjustment for alpha spending is not required.

#### Phase III Trial

From previous literature, we know that the survival of patients with cervical cancer with pelvic nodes is variable but ranges between 60% -80% at 5 years. In the literature use of extended field RT in populations at high risk for para-aortic nodal metastases was associated with significant improvement in the overall survival with reported hazard rates ranging between 0.5 - 0.8. For the current study we will assume that the 5 year overall survival is 70% in the control arm and that use of prophylactic EFRT will translate into an absolute improvement of 9% in the overall survival. This implies that the test arm will have a 5 year overall survival of about 79%. This corresponds to a hazard ratio of 0.75 which is a conservative estimate of the possible relative benefit of extended field radiotherapy. With a two sided type I error of 5% and a power of 80%, a total of 143 events is required to demonstrate an improvement in the overall survival corresponding to the hazard ratio of 0.75. This would need a total accrual duration of 5 years, and a minimum follow up duration of 4 years (such that the total trial duration of 9 years), a total of 462 patients (equal allocation) need to be accrued. Assuming a 15% loss to follow up, a total sample size of 530 patients is needed corresponding to an annual accrual of 106 patients.. An interim analysis for overall survival is not planned as the event rate is anticipated to be too low to prevent complete accrual into the study.

#### Accrual and duration of study

The estimated rate of accrual for this study is 5–7 patients a month. Thus, patient accrual is expected to be completed in 2 years. The total duration of the study is for 3 years, as the last recruited patient will be followed up for at least one more year to study the 3-year para aortic nodal recurrence rates in both the arms . The study will initiate recruitment in May 2024.

#### Allocation: sequence generation

Patients will be randomly allocated in the two arms in 1:1 ratio using the randomization module available in REDCap. Randomization sequence will be generated and stored in the RED Cap randomization module for random allocation of the patients. Once the screening is completed and the patient has given consent for the study, the random allocation will be done.The allocation will be done using the REDCap randomization module.

#### Allocation concealment mechanism

Randomization of the patient will be done using the randomization module implemented in the REDCap randomization module in the study database. Central randomization will be done to ensure allocation concealment.

#### Allocation Implementation

After screening and eligibility checks are completed, consenting patients will be randomized using the randomization module. The randomization will be done centrally at Tata Medical Center. RedCap provides a data access group feature which ensures that randomization is stratified by the institute. After filling the other stratification factors, the randomize button will be clicked which will then provide the arm allocation information. This information is permanently recorded in the database and cannot be altered in future.

### Blinding (masking)

Given the nature of the interventions proposed for this study, blinding will not be done.

### Procedure for unblinding if needed

As blinding is not being done in this trial, no unblinding will be required.

## Methods: Data collection, management and analysis

### Data collection plan

Data will be collected and maintained on a RedCap database maintained in Tata Medical Center. The randomization module of RedCap will be used for random allocation into the two arms. Participating centers will have access to the RedCap data entry system. The RedCap data forms can be printed out and used as such for clinical data entry in the trial if paper form entry is required.

The RedCap features for longitudinal data collection as well as repeating forms will be utilized for data collection. Planning CT and treatment plan data will be archived in an image bank called “CHAVI” which is developed in our institute [18].

### Plans to promote participant retention and complete follow-up

If a patient or investigator decides to stop the study treatment then the patient’s health status will be periodically reviewed via continued study visits or phone contact, or from their general practitioner or medical records to allow the collection of outcome data. Follow up assessments including completion of the quality of life questionnaires should still be completed if the patient is willing. In the event that a patient withdraws from the study entirely, the effective date of the notification will be the date on which their withdrawal is received by the study team. No information about the patient will be collected from that point in time onwards but any information collected prior to that date can be used and forms part of this study. For patients moving from the area during follow up, every effort should be made for the patient to be followed up at another participating trial centre and for that trial centre to take over responsibility for the patient. A copy of the patient Case Record Forms (CRF) will need to be provided to the new site after appropriate patient consent. Until the new centre agrees (in writing) to take over responsibility, the patient remains the responsibility of the original centre

### Data management

Access to the data will be available to the principal investigator of the study as well as to the co investigators after the completion of the study.The DSMC and trial statistician will have access to data while it is ongoing. The data quality and integrity will be checked at quarterly intervals using the data quality checking system available in Redcap. Patient identifiers will be noted as such in the Redcap database and the same will be used for ensuring that any data exports contain de-identified data only. Institutes will have access to the data of their own institute during the trial phase. Once the trial is completed, the main database will be checked for data consistency and quality and then closed for analysis. Additionally, the PI will be asked to sign off key CRFs electronically to ensure completeness and accuracy of the data. Source documents pertaining to the trial must be maintained by investigational sites. Source documents may include a subject’s medical records, hospital charts, clinic charts, the investigator’s subject study files and the results of diagnostic tests such as X-rays, laboratory tests, and electrocardiograms. All study-related documentation will be maintained for 10years following completion of the study or according to existing regulatory requirements.

#### Confidentiality

All study-related information will be stored securely on a trial specific REDCap database which has a password-protected access system and access to the information will be limited to only those that are part of the trial team. All the participant information that is stored in physical format will be in locked file cabinets in areas with limited access. The study data will be accessible to the local IRB for auditing, state and national regulatory bodies as necessary.For documents transferred between institutions, the data will be securely stored in the REDCap database and access restricted to the trial institute and the CTU through the use of the data access groups features in REDCap.

#### Plans for collection, laboratory evaluation and storage of biological specimens for genetic or molecular analysis in this trial/future use

Not planned for this study.

#### Statistical analysis for primary and secondary outcomes

Data collected in the dedicated study database in Redcap will be cleaned and cross checked by the investigating team at each center prior to the final analysis. A date cut off will be chosen which will be the date of closure of the database for the main analysis. This will usually be the date when the trial is after the required number of events (primary endpoint only) have been observed and will be discussed with the trial management team. Data will be analyzed using an established statistical package like R. The code or syntax for analysis will be saved in a file with required packages mentioned in the file for reproducible statistical analysis. REDCap automatically maintains a log of the file downloads for the analysis.

### Time to event endpoints

Median follow up will be calculated using the reverse kaplan meier method. For events like NRR, PNRR, LRR the cumulative incidence of the events will be computed considering death as a competing event. The Fine-Gray subdistribution hazard model will be used for computing the cumulative incidence of these events. The event rate at 5 years with the corresponding 95% intervals will also be computed using non-parametric Kaplan Meir method. Time to event endpoints like the overall survival, and progression free survival will be calculated using Kaplan Meier method and the unadjusted OS and PFS at 5 years, 95% confidence intervals of the estimate and the median values will be reported. Unadjusted and adjusted hazard ratios for OS and PFS will be estimated using the cox proportional hazard regression method. Adjustment variables will be:

1. Institute
2. Involvement of common iliac nodes: Present or Absent
3. Histology: Squamous or non-squamous
4. Number of pelvic nodes
5. Gross primary tumor volume (GTV) on MRI at diagnosis

Model assumptions will be checked for all models generated, and for cox regression will include the following:

1. For testing the proportional hazards assumption the scaled Schoenfeld residuals will be plotted against the log of time. The statistical test will be testing for a non-zero slope in a generalized linear regression of the scaled Schoenfeld residuals on the functions of time. If the proportional hazards assumption is not met then the accelerated failure time (AFT) model will be used. If the fit of the AFT model is not adequate, as demonstrated by a Q-Q plot, then a restricted mean survival time method as demonstrated by Royston & Parmar will be utilized(1). The restricted mean survival time approach is robust as it does not assume proportional hazards.
2. Influential observations or outliers will be checked using deviance residuals which are normalized transformations of the martingale residuals. The plot of these residuals against the observations should symmetrically distribute about zero with a standard deviation of 1.
3. Linearity assumptions for continuous covariates will be checked by plotting the Martingale residuals against the covariate. Fitted lines with loess function should appear linear to satisfy the linearity assumptions. If the same is found to be violated, continuous variables will be expanded by restricted cubic splines with 3 or more knots and will be used to model the continuous covariate.
4. In order to evaluate the model discrimination and calibration, bootstrapping with 500 bootstrap samples will be used to derive a calibration plot and optimism corrected C-index.

#### Additional statistical analysis

Pre-planned analysis of heterogeneity of treatment effect for the primary and secondary time to event endpoints will be done for the following subgroups:

1. Involvement of common iliac nodes: Present or Absent
2. Histology: Squamous or non-squamous
3. Common iliac nodal involvement : present or absent
4. Maximum primary tumor size: ≤ 4 cm or > 4 cm.

#### Methods in analysis to handle protocol non adherence and any statistical method to handle missing data

While every effort will be made to avoid missing data, a degree of missingness is anticipated. The trial sample size is calculated to account for a 5 % loss to follow up per year of the trial. However, if more than 10% of the population is lost to follow up then sensitivity analysis will be undertaken for the quality of life and adverse events endpoints.

Missing data for AEs and QoL will be analysed graphically stratified by arm and dropout time to evaluate if the missing data are Missing at Random or not. Sensitivity analysis will comprise of the following imputations to allow for MNAR conditions:

1. Best score is imputed for all
2. Worst score is imputed for all
3. Worst score imputed for the control arm and best score for the experimental arm
4. Best score imputed for the control arm and worst score for the experimental arm

The analyses will be repeated using these imputed datasets to generate sensitivity estimates of the QoL data. The above methodology provides a conservative estimate of the quality of life estimates. Additionally, another analysis will be conducted using complete cases only.

#### Plans to give access to the full protocol, participant level data and statistical code

The full protocol as well as the statistical analysis plan is available within the manuscript. There are no plans to make patient-level data available publically during the study recruitment, but we will consider the same once primary reporting of the outcomes is completed or as mandated by regulations.

#### Oversight and monitoring

##### Composition of the coordinating centre and trial steering committee

The trial will be managed through the CTU at the Tata Medical Center. All PIs of the collaborating institutes will be a part of the trial management committee. Study coordination, monitoring, data acquisition and management and statistical analysis will be performed by Tata Medical Center. Additionally, we will constitute an outcome assessment committee who will provide an independent and central assessment of the important trial outcomes.

#### Composition of the data monitoring committee, its role and reporting structure

The study will be monitored by the Institutional Data Safety Monitoring Sub-Committee, independent from investigators and a report will be submitted to the Institutional Ethics Committee (IEC). A continuing review application will be submitted by PI (primary investigator) at a regular interval (annually) to the IEC to continue the trial.

#### Adverse Events reporting

Events reporting all adverse events occurring during the study will be recorded using CTCAE V.5.0. The investigator at each site will be responsible for reporting the SAE to the respective Institutional Review Board as well as the PI at the CTU. Local IRB would be notified of any SAE within 24 hours on a working day or as per the institutional norm. SAEs must be reported up to 30 days from the end of the study intervention.

The following information should be provided for all SAEs:

1. Event description including classification according to NCI CTCAE
2. Primary and secondary diagnosis of the event (if death/hospitalization)
3. Severity/worst grade
4. Attribution to study intervention
5. Expectedness (listed in IB/product information)
6. Action taken with the study intervention
7. Impact of SAE (e.g. hospitalization details)
8. The outcome of SAE including end date if recovered

#### Frequency and plans for auditing trial conduct

Compliance to the protocol will be reviewed after accrual of the first 5, 25, 75 and 100 cases. Any protocol deviation needs to be notified to the respective IRB, to the chief investigator, coordinating centre and discussed in the trial management committee meetings. Additionally, PI at each centre should be notified about any changes made to the protocol in the interest of patient safety as a result of this—this will need updating in writing in the trial protocol with a change in the version number of the study protocol

#### Plans for communicating important protocol amendments to relevant parties (e.g. trial participants, ethical committees)

Changes and amendments to the protocol would only be made by the Trial Management Committee. Approval of amendments by the Institutional review board (IRB) would be required prior to their implementation. In some instances, an amendment may require a change to a consent form. In such cases, the form will be used only after approval from the IRB. All amendments will be stored in the sponsor site and will be shared with all participating sites through email as well as during regular trial management committee meetings.

#### Dissemination Policy

The results obtained from the study will be available to healthcare professionals and the public through open peer-reviewed scientific journals, conference presentations and abstracts.

#### Research ethics approval

The study has been approved by Tata Medical Center, Institutional Review Board on 20.06.2023(2021/TMC/240/IRB52) and PGIMER Chandigarh on 04.02.2023(PGI/IEC/2023/000037). The trial has been prospectively registered at the Clinical Trial Registry of India (CTRI) vide registration number: CTRI/2023/08/057075(30th August 2023)

### Informed consent

All patients will be provided with a patient information leaflet which will detail the study purpose, procedures, harms and expected benefits. Written informed consent will be obtained and signed by an independent witness as well as the PI or Co-PI or designated authorized person. The date, time and the name of the person obtaining the consent will be recorded in the medical records. Patients can withdraw consent at any time without ascribing a reason for the same. For patients who withdraw their consent for the trial, data up till the point of consent withdrawal will be retained but no further data will be collected. Informed consent forms are attached in Appendix 1.

### Trial status

The PROPARA trial was IRB approved and current protocol version is version 5.0 dated 13.01.2024. The trial was prospectively registered in the Clinical Trial Registry of India (CTRI) vide registration number: CTRI/2023/08/057075(30th August 2023).The trial will start recruiting patients from May 2024.

## Authors Contributions

The PROPARA Author group jointly drafted this manuscript. All authors read and approved the final manuscript. The list of authors in the author group is available in the Author details Table. All authors have contributed toward the drafting of the manuscript and formulation of the study protocol and have reviewed and approved the final version of the manuscript.

## Availability of data and materials

Data collected on patients will be made available based on regulatory norms after reporting of the primary outcomes.

## Consent for publication

Not applicable

## Ethics approval and consent to participate

The study has been approved by the Institutional Review Board of Tata Medical Center vide approval letter number 2021/TMC/240/IRB52 dated 10th October 2022. Additionally, IRB approval has been obtained from Post Graduate Institute of Medical Education and Research vide approval letter number PGI/IEC/2023/000037 dated 04/02/2023.All patients are provided with a patient information sheet translated into the vernacular language and written informed consent is obtained before accrual.

## Appendix 1 English Consent

### INTRODUCTION

You are invited to participate in this study because you have been diagnosed with cervical cancer that has spread to the lymph nodes in your lower abdomen. This type of cancer is what we call stage III cancer (Stage IIIC1). To treat this disease, your doctor has advised radiotherapy and chemotherapy. With this study we want to understand if we can improve the chances of a cure if we treat your upper abdominal lymph nodes along with the lower abdomen with radiotherapy.

Before you participate, we would like to explain the purpose of this study and give you the opportunity to ask questions. Read the information provided here carefully.

Participation in this study is completely voluntary. If you don’t want to participate, you don’t have to. Whether you participate or not you will get the right treatment.

If you agree to participate, please sign the informed consent form. You will be given a copy of this document to take home with you.

### WHAT IS THE PURPOSE OF THIS RESEARCH?

When cervical cancer has spread to the lymph nodes in the lower abdomen, it is treated with radiation and chemotherapy. After giving radiation from outside, radiation from inside is also given which we call brachytherapy. We know that despite this treatment, about 10-15 out of 100 women will have the disease coming back or spreading to the upper abdominal lymph nodes.

The present research is attempting to see whether the addition of prophylactic para aortic radiation therapy to pelvic radiotherapy will help in reducing the chance of para aortic failure and improving the overall survival. This may help the patient maintain their quality of life and help them to survive longer .

#### The study plans to include about 385 patients with stage IIIC1 cervical cancer

### WHAT DOES PARTICIPATION IN THE STUDY INVOLVE?

In this study we will divide the treated women into two groups. One group (group A) will be treated with external radiation to the pelvis or lower abdomen. The other group (group B) will be treated with radiation to the lower abdomen and upper abdominal lymph nodes. Besides this, all other treatments (chemotherapy and brachytherapy) will be done in the same manner in both groups.

If you agree to participate in this study, you will be assigned to a treatment group through a process called randomization. Randomization is like a lottery. Neither you nor your doctor can change the outcome of this randomisation. Randomization will be done impartially by a central computer.

Additional chemotherapy may be given before radiotherapy as per the institute’s protocol. If chemotherapy is given before radiotherapy, you will have two chemotherapy medicines given into your vein (palitaxel and carboplatin). These chemotherapy injections will be given to you 6 times (once a week). Apart from this, you will also be given other medicines along with the injection to control nausea, vomiting and allergic reactions caused by chemotherapy. This chemotherapy is not part of this research.

#### Investigations before treatment

Physical examination, general blood tests, CT scan of chest and abdomen and MRI scan of lower abdomen will be done before the treatment. This research is generally done before this treatment and no new tests will be done for the research.

#### Radiation Therapy

For radiation therapy, you will be called to plan radiation treatment. This will involve undergoing a CT scan which will be done to accurately plan the radiotherapy. The radiotherapy will be given with the best possible and most advanced technique in both groups to decrease possible side effects. The duration of external beam radiotherapy will be around 5 weeks. The duration will be explained to you by your trial doctor.

#### Follow Up

Your condition will be monitored regularly during and after your treatment. You will need to visit the hospital every three months for the first 2 years after treatment. You will have to go to the hospital every 6 months after two years. After 5 years you will have to come once a year.

You will have an MRI scan of your abdomen 3 months after your treatment is completed. Apart from this, you will also be asked to undergo other tests if you have any discomfort later. This scan or test will be done in the same way in both groups and these tests are part of the normal treatment. You do not need to do any new test to take part in the study.

##### Investigations after treatment

During the follow-up, you will be physically examined by your physician. MRI pelvis will be done before your brachytherapy or internal radiation procedures after 3 months after completion of your treatment. There are no additional costs associated with participating in this research project, nor will you be paid.

##### You will have to pay for non-study medicines that are given as routine treatment for your condition according to hospital policy

### WHAT ARE THE ALTERNATIVES TO PARTICIPATION?

You do not have to participate in this research project to receive treatment at this hospital. If you decide not to take part in this study, you will be given the usual standard treatment (abdominal radiation, chemotherapy and brachytherapy). Also you can get your treatment in other hospital if you want. You can also get a second opinion about your treatment.

### WHAT ARE THE POSSIBLE RISKS OF PARTICIPATING?

Medical treatments often cause side effects. You may experience some or all of the side effects that the doctor involved in this study told you about. These side effects can be mild, moderate or severe. There may also be side effects that we did not expect or know about. These side effects can be serious. Tell your doctor about any new or unusual symptoms you experience. Also, in case of physical discomfort, immediately come to the emergency department of the hospital. You can come to the emergency department any time of the day or night and you do not need to make an appointment.

We have informed you about the possible side effects of the treatment. Common side effects are listed below. Most of the side effects during treatment go away after the treatment ends. But sometimes these side effects can be severe, chronic or permanent. If a serious side effect or reaction occurs, your study doctor may need to stop your treatment. Your study doctor will discuss with you the best way to manage any side effects.

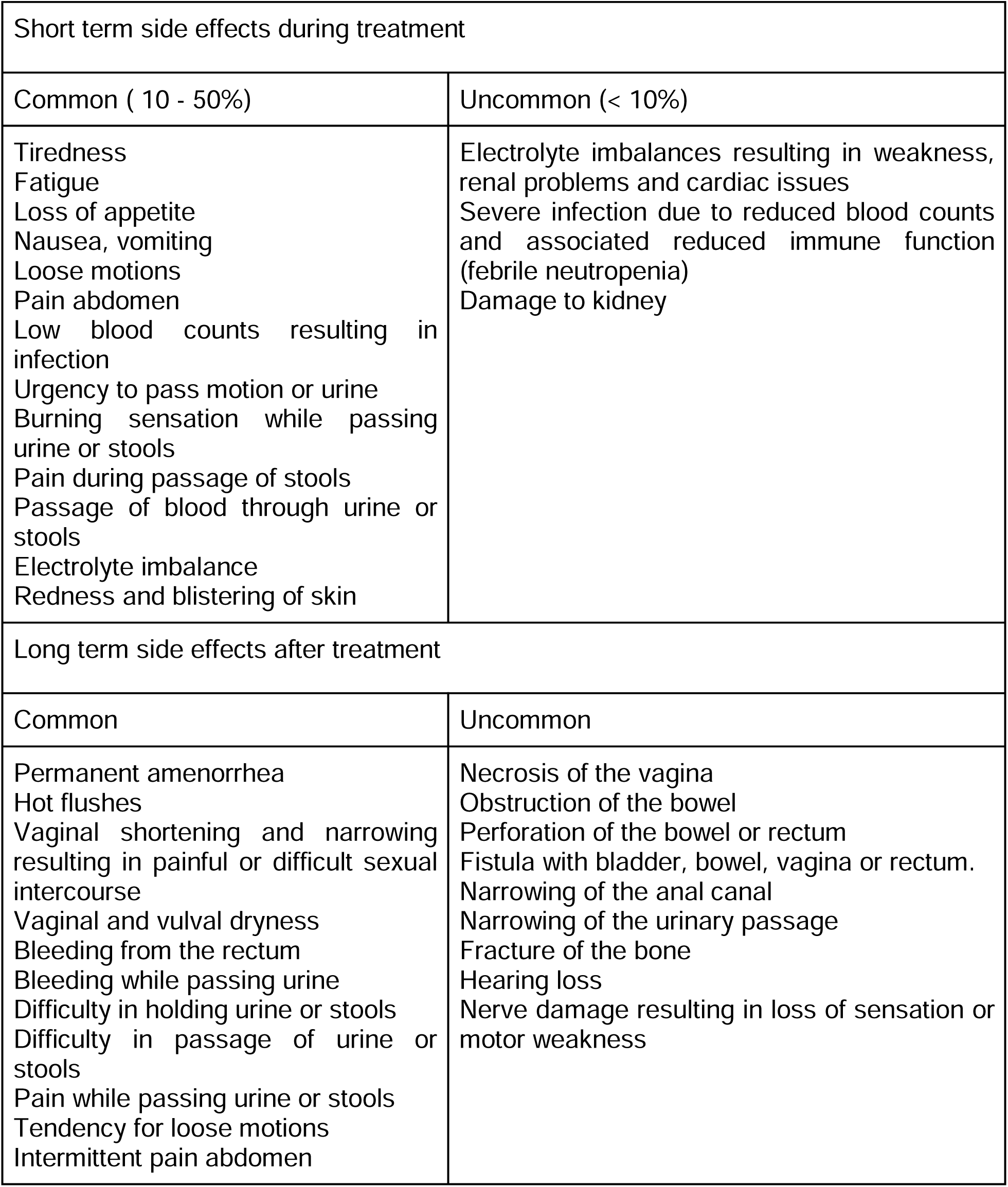

| Short term side effects during treatment |  |
| --- | --- |
| Common ( 10 - 50%) | Uncommon (< 10%) |
| Tiredness<br>Fatigue<br>Loss of appetite<br>Nausea, vomiting<br>Loose motions<br>Pain abdomen<br>Low blood counts resulting in infection<br>Urgency to pass motion or urine<br>Burning sensation while passing urine or stools<br>Pain during passage of stools<br>Passage of blood through urine or stools<br>Electrolyte imbalance<br>Redness and blistering of skin | Electrolyte imbalances resulting in weakness, renal problems and cardiac issues<br>Severe infection due to reduced blood counts and associated reduced immune function (febrile neutropenia)<br>Damage to kidney |
| Long term side effects after treatment |  |
| Common | Uncommon |
| Permanent amenorrhea<br>Hot flushes<br>Vaginal shortening and narrowing resulting in painful or difficult sexual intercourse<br>Vaginal and vulval dryness<br>Bleeding from the rectum<br>Bleeding while passing urine<br>Difficulty in holding urine or stools<br>Difficulty in passage of urine or stools<br>Pain while passing urine or stools<br>Tendency for loose motions<br>Intermittent pain abdomen | Necrosis of the vagina<br>Obstruction of the bowel<br>Perforation of the bowel or rectum<br>Fistula with bladder, bowel, vagina or rectum.<br>Narrowing of the anal canal<br>Narrowing of the urinary passage<br>Fracture of the bone<br>Hearing loss<br>Nerve damage resulting in loss of sensation or motor weakness |

Because chemotherapy and radiation are given to all patients, all patients may experience some or all of these side effects. We know that upper abdominal radiotherapy treatment may increase the risk of some side effects.

### WHAT ARE THE COSTS OF PARTICIPATING IN THIS TRIAL?

No different treatments or tests are being performed in this study. There is no additional cost to you for radiation treatment of the upper abdominal lymph nodes. You will pay the usual cost of radiotherapy.

### WILL THERE BE REIMBURSEMENT FOR PARTICIPATION IN THE TRIAL?

No financial reimbursement is planned for participation in the study.

### WHAT ARE THE POTENTIAL BENEFITS OF PARTICIPATING IN THE TRIAL?

There may be no direct benefits to you due to participation in the trial. You will receive quality treatment as per standard guidelines in both arms. There is no assurance however that you will get benefit from this study.

However, your participation will contribute to medical knowledge about the best way to treat patients with diseases like yours. Please remember that many of the most effective treatments used today are the result of clinical trials done in the past.

### CONFIDENTIALITY OF STUDY AND MEDICAL RECORDS

- All identifying information will be kept confidential
- Data will be securely stored in a database in Tata Medical Center Kolkata and PGIMER Chandigarh
- Your information will be identified by a code number which can be only accessed by the research team
- Your personal information will not be disclosed without prior permission or as required by law
- Anonymized data will be used for analysis

### COMPENSATION FOR PROTOCOL RELATED INJURY

- If you suffer physical injury as a result of taking the test treatment you are eligible to receive financial compensation.
- If you die as a result of taking the test treatment then your dependents are eligible to receive compensation.
- Compensation will be given after approval of the Institutional Ethics Committee (IEC).
- If you suffer any physical harm due to the research treatment, you will be treated free of charge. This will be given for any injury caused during your treatment and up to 30 days after your radiation treatment is completed.
- If the Institutional Ethics Committee (IEC) determines that your injury is not related to the research, you will have to bear the cost of your treatment.
- brachytherapy or other treatment) you have to bear the cost yourself.
- Any medical treatment taken outside TMC will be covered only if you inform us immediately after the admission or at the maximum, earliest possible time of the next day (if admission was too late at night).
- This will be provided till the time it is established that the injury is not related to the clinical trial.

### PARTICIPATION

Your participation in this study is voluntary; You may refuse to participate at any time without penalty and without loss or benefit.

If you withdraw your consent before the study is completed, you will receive the usual standard care for your disease. Your participation will not adversely affect your subsequent treatment or relationship with your doctor. If you withdraw consent from the study before data collection is complete, your information after the date of withdrawal will not be recorded in the study report.

### WHOM TO CONTACT IF YOU HAVE QUESTIONS

If you have questions at any time about the study or the procedures, (or you experience adverse effects as a result of participating in this study,) you may contact the researcher.

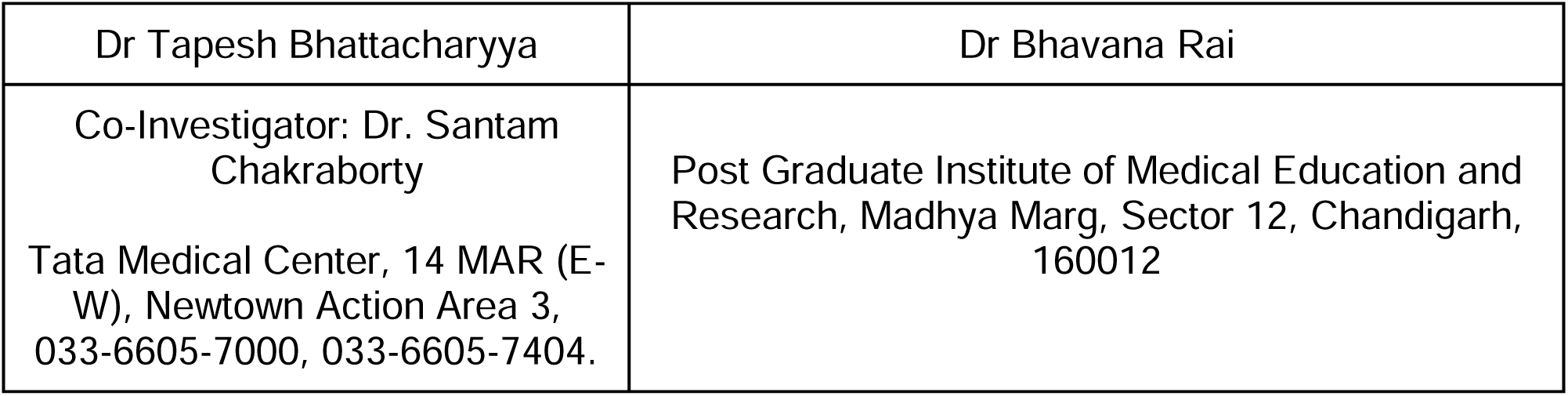

Tata Medical Center Institutional Review Board (TMC-IRB) can be contacted on 03366057579 if you have any concerns about the project. The director of TMC is the appellate authority in case you have issues with the IRB decision on your concern.

### INFORMED CONSENT FORM TO PARTICIPATE IN THE CLINICAL TRIAL

1. I understand that I am being invited to take part in the research study. I confirm that I have read and understood the information sheet dated ***<u>Version 2.0, Dated: 07/01/2024</u>*** for the above study and have had the opportunity to ask questions.
2. I understand that my participation in the study is voluntary and that I am free to withdraw at any time, without giving any reason, without my medical care or legal rights being affected.
3. I understand the risks and potential benefits of this research study that were explained to me. I freely give my consent to take part in the research study described in this form.
4. I understand that institutional review boards and regulatory authorities may access my health records for the current study and any subsequent research related to it without my permission. This may also be after I have withdrawn my consent. I understand that my identity will not be disclosed in any information disclosed or disclosed to third parties.
5. I agree not to restrict the use of any data or results that arise from this study provided such use is only for the scientific purpose(s).
6. I have read the above information and agreed to participate in this study. I have received a copy of this form.

**Patient**

Signed:

Print Name:

Date:

**Witness** Signed:

Print Name:

Date:

**Investigator**

Signed:

Print Name:

Date:

